# Physician-confirmed and administrative definitions of stroke in UK Biobank reflect the same underlying genetic trait

**DOI:** 10.1101/2021.09.30.21264348

**Authors:** Kristiina Rannikmäe, Konrad Rawlik, Amy C Ferguson, Nikos Avramidis, Muchen Jiang, Nicola Pirastu, Xia Shen, Emma Davidson, Rebecca Woodfield, Rainer Malik, Martin Dichgans, Albert Tenesa, Cathie Sudlow

## Abstract

**Background:** Stroke in UK Biobank (UKB) is ascertained via linkages to coded administrative datasets and self-report. We studied the accuracy of these codes using genetic validation.

**Methods:** We compiled stroke-specific and broad cerebrovascular disease (CVD) code lists (Read V2/V3, ICD-9/-10) for medical settings (hospital, death record, primary care) and self-report. Among 408,210 UKB participants we identified all with a relevant code, creating 12 stroke definitions based on the code type and source. We performed genome-wide association studies (GWASs) for each definition, comparing summary results against the largest published stroke GWAS (MEGASTROKE), assessing genetic correlations, and replicating 32 stroke-associated loci.

**Results:** Stroke case numbers identified varied widely from 3,976 (primary care stroke-specific codes) to 19,449 (all codes, all sources). All 12 UKB stroke definitions were significantly correlated with the MEGASTROKE summary GWAS results (rg 0.81-1) and each other (rg 0.4-1). However, Bonferroni-corrected confidence intervals were wide, suggesting limited precision of some results. Six previously reported stroke-associated loci were replicated using ≥1 UKB stroke definitions.

**Conclusions:** Stroke case numbers in UKB depend on the code source and type used, with a 5-fold difference in the maximum case-sample size. All stroke definitions are significantly genetically correlated with the largest stroke GWAS to date.

## Introduction

UK Biobank (UKB) is a prospective population-based cohort study with extensive phenotype and genotype information on >500,000 participants from England, Scotland and Wales (www.ukbiobank.ac.uk). It is an open access resource, established to facilitate research into the determinants of a wide range of health outcomes, particularly those relevant in middle and older age (Sudlow, 2015). An example of such a disease is stroke, the second most common cause of death worldwide and a major global cause of disability (Lozano, 2012).

Disease outcomes in UKB are ascertained chiefly via linkages to routinely collected, coded, national administrative health datasets. In addition, data on self-reported medical conditions was collected at recruitment. However, to use these data appropriately, researchers need to select which particular disease codes to use for their study and have an understanding of their accuracy. For example, to identify stroke cases, existing codes can be divided into those that are stroke-specific and those that fall under the broad cerebrovascular disease (CVD) category. Stroke-specific codes are used to code acute stroke events where the clinician is confident about the diagnosis and can usually assign a subtype. In contrast, broad CVD codes also capture cases with: (i) phenotypes which pose a high risk for a subsequent stroke (e.g. a code for transient ischaemic attack, an unruptured aneurysm, or carotid artery stenosis); (ii) a past history of stroke with residual symptoms (e.g. a code for sequelae of cerebral infarction); (iii) events where there may be some diagnostic uncertainty (e.g. a code for unspecified cerebrovascular disease); and (iv) intracranial haemorrhages other than intracerebral or subarachnoid haemorrhage (e.g., extradural or subdural haemorrhages, which most clinicians consider different from stroke). Including codes from the broad CVD category will therefore significantly increase the overall number of cases identified, but while this is likely to include at least some misclassified true acute stroke cases, non-stroke cases will also be included.

In a systematic review of studies validating stroke code accuracy from case-note review, the overall positive predictive value (proportion of true-positive cases among all identified cases) for identifying acute stroke cases was consistently >70% for stroke-specific codes, dropping to <50% in many studies when broad CVD codes were included (Woodfield, 2015 & Rannikmäe, 2020). For self-reported stroke events, the positive predictive value ranged from 22% to 87% across different studies, making it hard to draw firm conclusions (Woodfield, 2015). While case-note review for code validation is often considered a gold-standard, this method also has its limitations. It is time-consuming and labour-intensive, so can only be achieved in relatively small numbers of cases, with limited precision of the results. In addition, the results rely on: (i) accessing the complete relevant medical record; (ii) the detail and quality of the medical record; (iii) the qualification of the person reviewing the notes; (iv) the inter-adjudicator agreement, which we know is not perfect even between highly specialised clinicians; and (v) the consistency of results across different healthcare settings/providers (Liberman, 2018).

We set out to supplement current knowledge about the accuracy of stroke codes with a method making use of large-scale genetic data, which we refer to as ‘genetic validation’. The fundamental idea is to use existing knowledge of genetic associations with a disease (in this case acute stroke), to assess how well various potential code lists capture people who truly have this disease, which in turn could be used to harmonise disease definitions across cohorts and health systems (Manolio, 2020). If the code list captures true-positive cases, we would expect the genetic associations that result from stroke cases identified through coded data to closely mirror the genetic association results from previous studies of stroke.

## Methods

### Study setting

We included all 408,210 UKB white British ancestry participants in this study. We restricted our analyses to this ancestry subgroup because it covers 94% of the UKB participants and allowed us to achieve a good balance between attaining sufficient case numbers while reducing population stratification and analytic complexity. As part of the UK Biobank recruitment process, informed consent was obtained from all individual participants included in the study. At the time of the study, UKB had linked hospital admissions and death registry administrative coded data available for all participants, and primary care administrative coded data for 47% of the cohort (191,146), covering the time period up to March and September 2019, respectively (S1 Table). In addition, all participants self-reported pre-existing health conditions during an interview at recruitment. The subset of the cohort with primary care data available was similar to the whole cohort with respect to age at recruitment, sex and Townsend deprivation index (S2 Table).

### Identifying stroke cases in UKB

We compiled stroke-specific and broad cerebrovascular disease (CVD) code lists for each medical setting (hospital admission, death record, primary care) and self-report. This process was informed by previously published codes where available (Woodfield, 2015 & Rannikmäe, 2020), supplemented by the selection of additional codes by expert clinicians (authors KR, CLMS, ED, RW) on discussion and mutual agreement. This resulted in a total of 8 code lists, covering the ICD-9/ICD-10, Read Version 2, Clinical Terms Version 3 (Read Version 3) and UKB self-report illness coding systems (S3 Table).

Next, we identified all participants with a relevant code from any of the code lists and created 12 different ways of defining stroke cases in UKB based on the code type (stroke-specific, broad CVD) and source (hospital admission, death record, primary care, self-report). This resulted in 12 partially overlapping case-control groups, where cases were all the individuals with a stroke code for the particular stroke definition, and all the remaining participants acted as controls. A specific UKB participant could therefore be a stroke case for one definition and a control for another definition.

### Genome-Wide Association Studies

We performed 12 genome-wide association studies (GWASs), one for each case-control set (i.e. for each definition of stroke cases and their controls). We applied a linear mixed model method using the BoltLMM software package (v2.3.4) software (Loh, 2015). We included the following as covariates: genotyping array, UKB assessment centre, sex, age at recruitment, and principal components one to ten. We filtered the results for single nucleotide polymorphisms (SNPs) with an imputation quality INFO score ≥0.9 and minor allele frequency ≥1%. After filtering the results for SNP imputation quality and minor allele frequency, we included 9,524,428 SNPs. We converted the linear mixed model effects to odds ratios using R code provided in https://shiny.cnsgenomics.com/LMOR/ (Lloyd-Jones, 2018).

### Analyses of GWAS results

We compared summary results from our 12 GWASs against the largest published stroke GWAS meta-analysis project - the MEGASTROKE study. The MEGASTROKE study is a meta-analysis of 29 stroke GWASs (17 including individuals of European ancestry) and does not include UKB data. Almost all studies included in MEGASTROKE (covering >95% included cases) required the stroke diagnosis to be confirmed by a medical professional or required evidence of stroke from >1 source, even if the initial case ascertainment included using administrative codes (Malik, 2018). All analyses were done using R software version 3.6.2.

#### Genetic correlation with the MEGASTROKE study results

We applied a high-definition likelihood method using the HDL software (Ning, 2020) to assess the genetic correlation between our GWAS results using the 12 stroke definitions, and the MEGASTROKE study GWAS summary results for any stroke subtype in European samples. Genetic correlation (rg) is the proportion of variance that two stroke definitions share due to genetic causes. A genetic correlation of 0 implies that the genetic effects on one definition are independent of the other, while a correlation of 1 implies that all of the genetic influences on the two definitions are identical. We assessed if the correlation was significantly different from 0 and from 1, setting the p-value significance threshold to 0.0042 after a Bonferroni correction for the 12 tests. We displayed the results (correlation measured as rg) on a heatmap. We also display Bonferroni corrected confidence intervals to aid interpretation.

#### Genetic correlation within our study definitions

We then used the HDL software to assess genetic correlations within our study across the 12 definitions. We set the significance threshold to 0.0024 after a Bonferroni correction for 7 independent non-overlapping case-control definitions (definitions not in bold in Table 1), resulting in 21 correlation tests. We also display Bonferroni corrected confidence intervals to aid interpretation.

**Table 1.** Number and demographic characteristics of stroke cases identified in UKB.

| **STROKE DEFINITION** | **NUMBER OF CASES** | **NUMBER OF CONTROLS** | **Mean (median) age at recruitment (years)** | **Mean (median) age; age range at stroke (years)** | **Sex (% female)** |
| --- | --- | --- | --- | --- | --- |
| Stroke-specific code from hospital/death records | 6,887 | 401,323 | 61 (63) | 63 (64); 31 to 79 | 40 |
| Stroke-specific code from primary care | 3,976 | 404,234 | 61 (63) | 59 (61); 1 to 79 | 41 |
| **Stroke-specific code from any medical setting** | **8,665** | **399,545** | 61 (63) | 61 (63); 1 to 79 | 41 |
| Broad CVD code from hospital/death records | 5,725 | 402,485 | 62 (63) | 64 (65); 31 to 79 | 43 |
| Broad CVD code from primary care | 4003 | 404,207 | 62 (63) | 62 (63); 1 to 79 | 41 |
| **Broad CVD code from any medical setting** | **8,085** | **400,125** | 62 (63) | 63 (64); 1 to 79 | 44 |
| Stroke-specific or broad CVD code from hospital/death records | 12,612 | 395,598 | 61 (63) | 63 (64); 31 to 79 | 41 |
| Stroke-specific or broad CVD code from primary care | 7,979 | 400,231 | 62 (63) | 60 (62); 1 to 79 | 41 |
| **Stroke-specific or broad CVD code from any medical setting** | **16,750** | **391,460** | 62 (63) | 62 (63); 1 to 79 | 42 |
| Specific self-reported stroke event | 5,915 | 402,295 | 61 (62) | 53 (55); 0 to 70 | 41 |
| **Specific or non-specific self-reported stroke event** | **7,536** | **400,674** | 61 (63) | 53 (55); 0 to 70 | 42 |
| **Any code or self-reported event** | **19,449** | **388,761** | 61 (63) | 60 (61); 0 to 79 | 43 |
| Across all UKB participants | 408,210 |  | 57 (58) | Not applicable | 54 |

We considered code and self-reported event age missing if it predated the date of birth, contained a negative value, or was recorded by UKB as unreliable or missing – this affected 0.3% to 2.1% cases depending on the stroke definition. CVD: cerebrovascular disease;

#### Replicating the MEGASTROKE study stroke-significant loci

The MEGASTROKE study identified 32 genetic loci significantly associated with stroke. We identified these loci (the lead SNP for each locus) in our GWAS summary results and considered a locus to be replicated (i.e. also significantly associated with the respective stroke definition in our data) if the p-value of association in our GWAS was <0.00156 (Bonferroni corrected for 32 loci). We compared the number of replicated loci across our summary definitions. We compared the effect sizes of the associations between MEGASTROKE trans-ethnic and European ancestry GWASs and our GWAS summary results. Where the lead SNP was not available in our data, we identified SNPs in moderate LD (r^2^ > 0.7 in the 1000 Genomes GBR population using the Ensembl LD calculator https://www.ensembl.org/Homo_sapiens/Tools/LD) with the lead SNP, and if any SNPs in LD available in our data were identified, we examined their associations instead. We displayed results for five of our summary definitions of stroke cases and their controls: stroke-specific code from any medical setting; broad CVD code from any medical setting; stroke-specific or broad CVD code from any medical setting; specific or non-specific self-reported stroke event; any code or self-reported event. We highlighted significantly associated (i.e. replicated) loci.

We also calculated our expected power to replicate the 32 loci using the Genetic Association Study (GAS) Power Calculator (http://csg.sph.umich.edu/abecasis/gas_power_calculator/index.html), assuming a stroke prevalence of 2.26% and inputting the disease allele frequency and genotype relative risk estimates from the MEGASTROKE publication Table 1 (Malik, 2018).

## Results

### Stroke cases in UKB

The number of relevant codes for identifying stroke cases varied widely depending on the coding system (ICD versus Read versus self-report) and code type (stroke-specific versus broad CVD code) – from less than five codes for a specific self-reported stroke event, to >500 codes when including all possible codes across all coding systems. The stroke-specific and broad cerebrovascular disease (CVD) code lists for each medical setting and self-report are shown in S3 Table.

The number of stroke cases identified among the 408,210 participants also varied widely depending on the code type and source used – from 3,976 cases in primary care when using stroke-specific codes, to 19,449 cases when including all possible code combinations (stroke-specific and broad CVD) across all sources (hospital admission, death record, primary care, self-report) (Table1).

The code source for cases with a stroke-specific code was: self-report only in 27%, primary care only for 9%, hospital/death record code only for 29%, and >1 source for 35% (S1 Figure). The code source for cases with either a stroke-specific or a broad CVD code was: self-report only in 14%, primary care only for 15%, hospital/death record code only for 34%, and >1 source for 37% (S1 Figure). These proportions are calculated based on the primary care data being currently available only for ∼50% of the participants, and so will change when primary care data for the whole cohort become available.

The overall proportion of prevalent codes (i.e. first code predates participant’s recruitment to UKB) versus incident codes (i.e. first code date occurs after participant’s recruitment to UKB) was the same for stroke-specific and broad CVD categories: 38% prevalent versus 62% incident codes. These proportions are dependent on the updates to different linked health datasets and the proportion of incident codes will continue to increase with increasing duration of follow up. (S1 Table).

Mean and median age at recruitment were higher among stroke cases (for all stroke definitions) than for the whole cohort of UKB participants (mean age 61 to 62 years vs 57 years; median age 62 to 63 years vs 58 years). Mean and median age at the time of stroke (in case of multiple events, age at the earliest event was taken) was higher for coded diagnoses from the medical setting compared to self-reported events (mean age 62 years vs 53 years, median age 63 years vs 55 years, respectively). This is to be expected, considering that all self-reported events were recorded at the time of recruitment, whereas medical codes also capture diagnoses after recruitment during follow up. The proportion of women was lower among stroke cases than across all UKB participants (43% for those with any medical setting or self-reported code vs 54% for all UKB). This is to be expected as age-specific incidence rates are substantially lower in women than men in younger and middle-age groups, but these differences narrow down so that in the oldest age groups, incidence rates in women are approximately equal to or even higher than in men (Virani, 2020). (Table 1).

### Analyses of GWAS results

#### Genetic correlation with the MEGASTROKE study results

All 12 UKB stroke definitions were significantly correlated with the MEGASTROKE summary GWAS results, with genetic correlations (rg) ranging from 0.81 to 1, and confidence intervals overlapping. The p-values for difference from 1 were not significant, compatible with perfect correlation. However, the Bonferroni corrected CIs were wide, especially for 5 of the 12 tests, where the lower confidence limit was <0.7, limiting the precision of some of these results (Figure 1, Figure 2, S4 Table).

**Figure 1.**
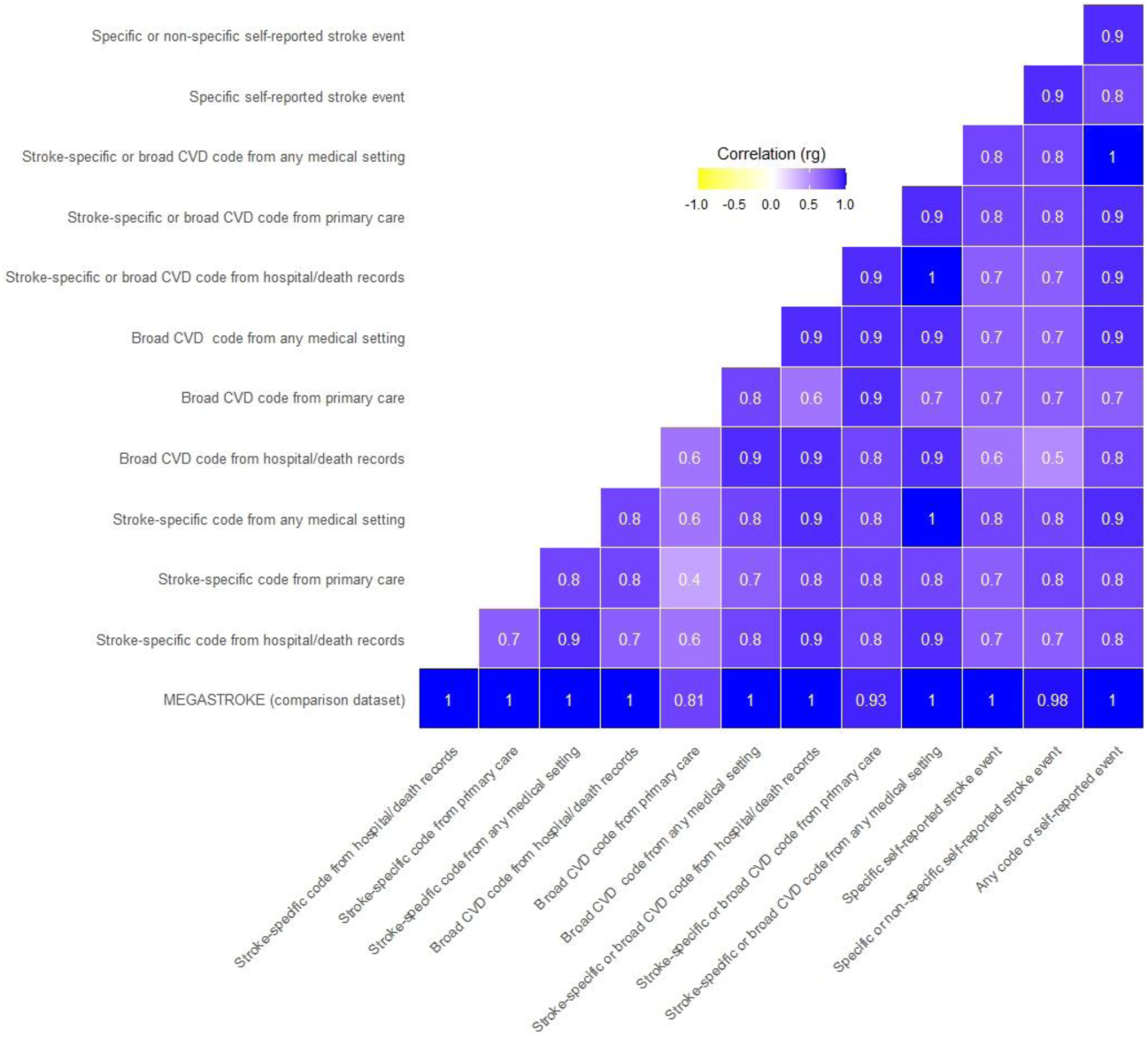
Genetic correlation of UKB stroke definitions with MEGASTROKE and each other. Legend: Where the rg was >1, we rounded it to 1.

**Figure 2.**
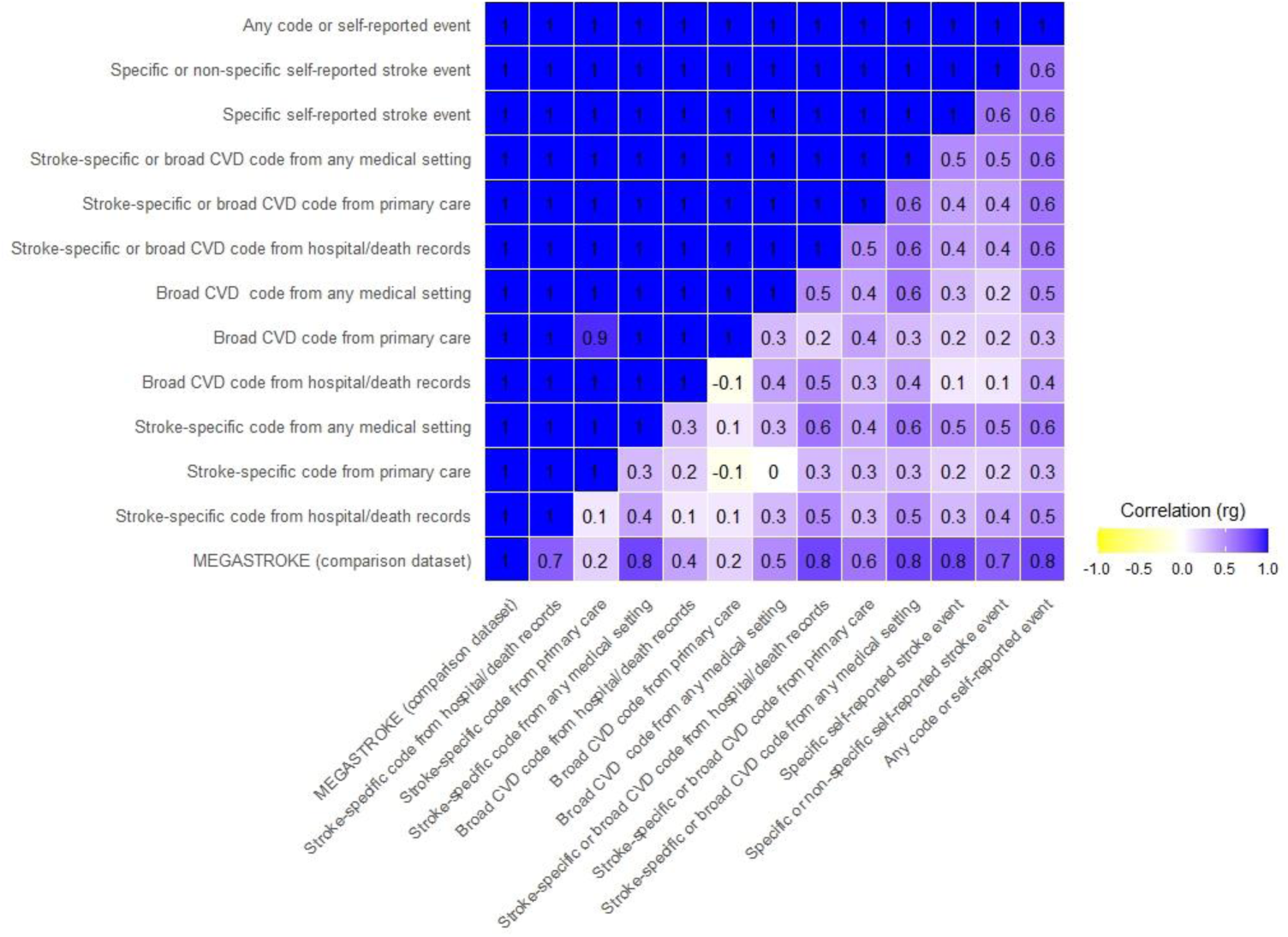
Confidence intervals of genetic correlation of UKB stroke definitions with MEGASTROKE and each other. Legend: The upper triangle displays Bonferroni corrected upper confidence intervals, and the lower triangle displays Bonferroni corrected lower confidence intervals. Where the rg was >1, we rounded it to 1.

#### Genetic correlation within our study definitions

The UKB summary definitions in our study were all significantly correlated with each other (all p-values significantly different from 0), with rg ranging from 0.4 to 1. Again, the Bonferroni corrected CIs were wide, with the lower confidence limit even suggesting the possibility of a negative correlation for two comparisons. (Figure 1, Figure 2, S5 Table).

#### Replicating the MEGASTROKE study stroke-significant loci

Within our GWASs, 6 of the 32 previously reported stroke-associated loci were replicated by one or more definitions. Analyses using stroke-specific codes and analyses using any code or self-reported event both replicated the biggest number of known stroke loci (5 of 32). The power from additional cases for the latter category did not result in replicating more loci than stroke-specific codes alone. However, for three of the five replicated loci, the p-values were smaller in the larger dataset (analyses using any code or self-reported event) suggesting a more robust replication when using the broadest definition of stroke in UKB. Within our data, effect sizes (expressed as odds ratios) were similar across the stroke definitions, with overlapping Bonferroni corrected confidence intervals.

For two of the six replicated loci (*PITX2* and *HDAC9–TWIST1*), the effect size of the association (odds ratio) was bigger in the MEGASTROKE dataset than in our data (across all five summary stroke definitions). These two loci are known to be associated with particular stroke subtypes – *PITX2* with cardioembolic and *HDAC9–TWIST1* with large artery stroke [10]. (Figure 3, Table 2, S6 Table).

**Figure 3.**
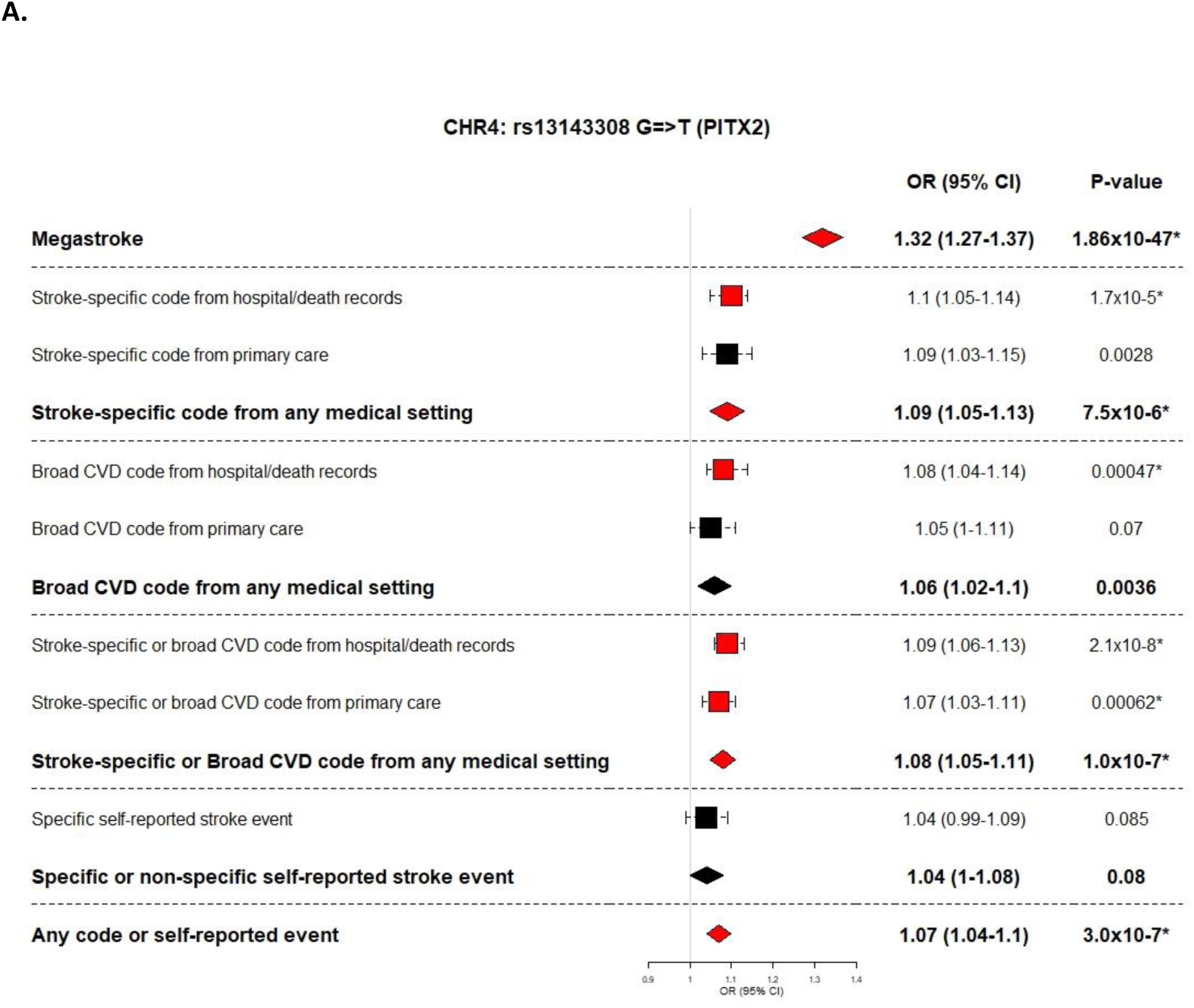
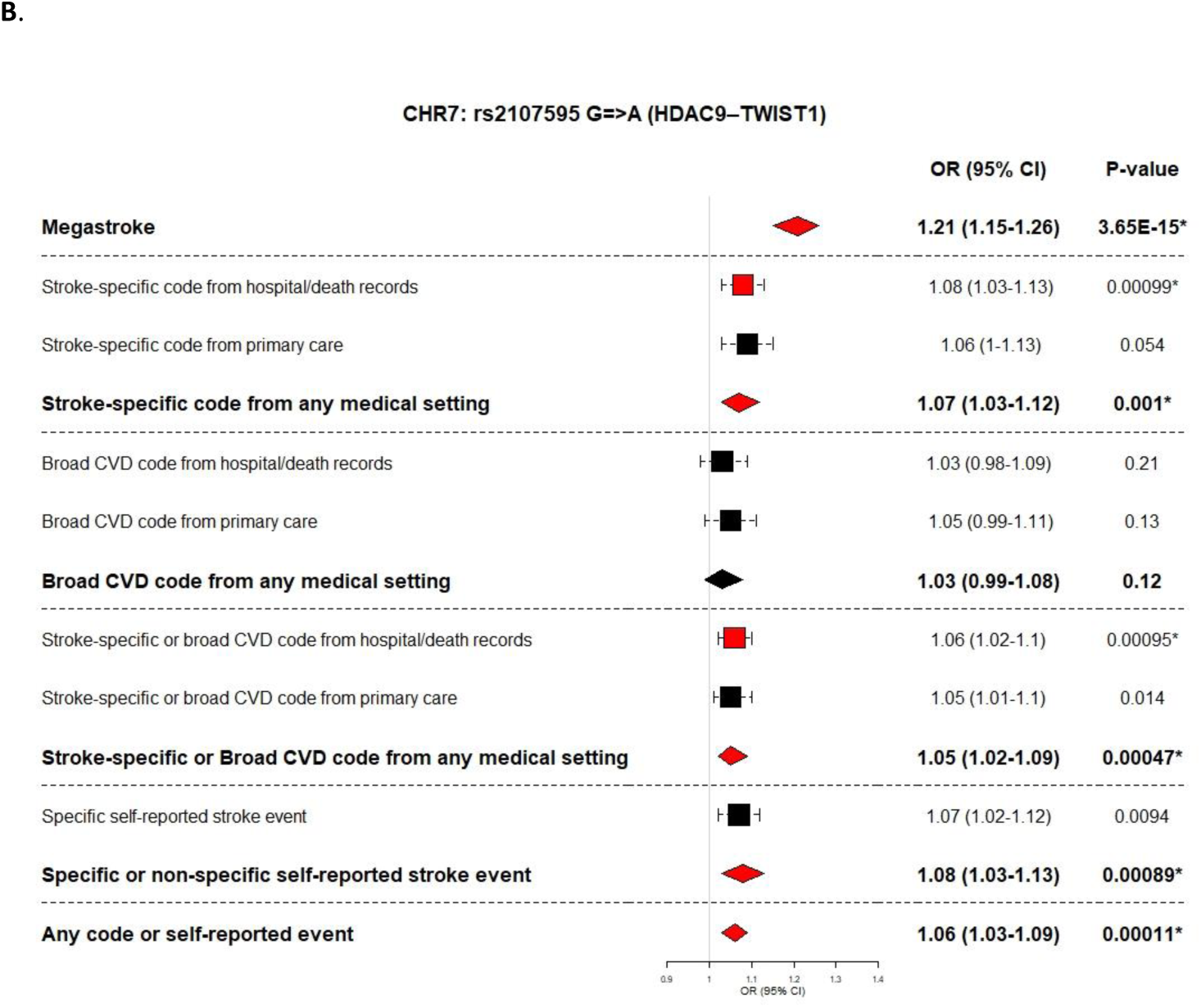
Megastroke stroke subtype-significant loci replicated using UK Biobank stroke definitions. Legend: MEGASTROKE odds ratio and p-value is shown for the analyses (European or trans-ethnic) showing the lowest p-value

**Table 2.**
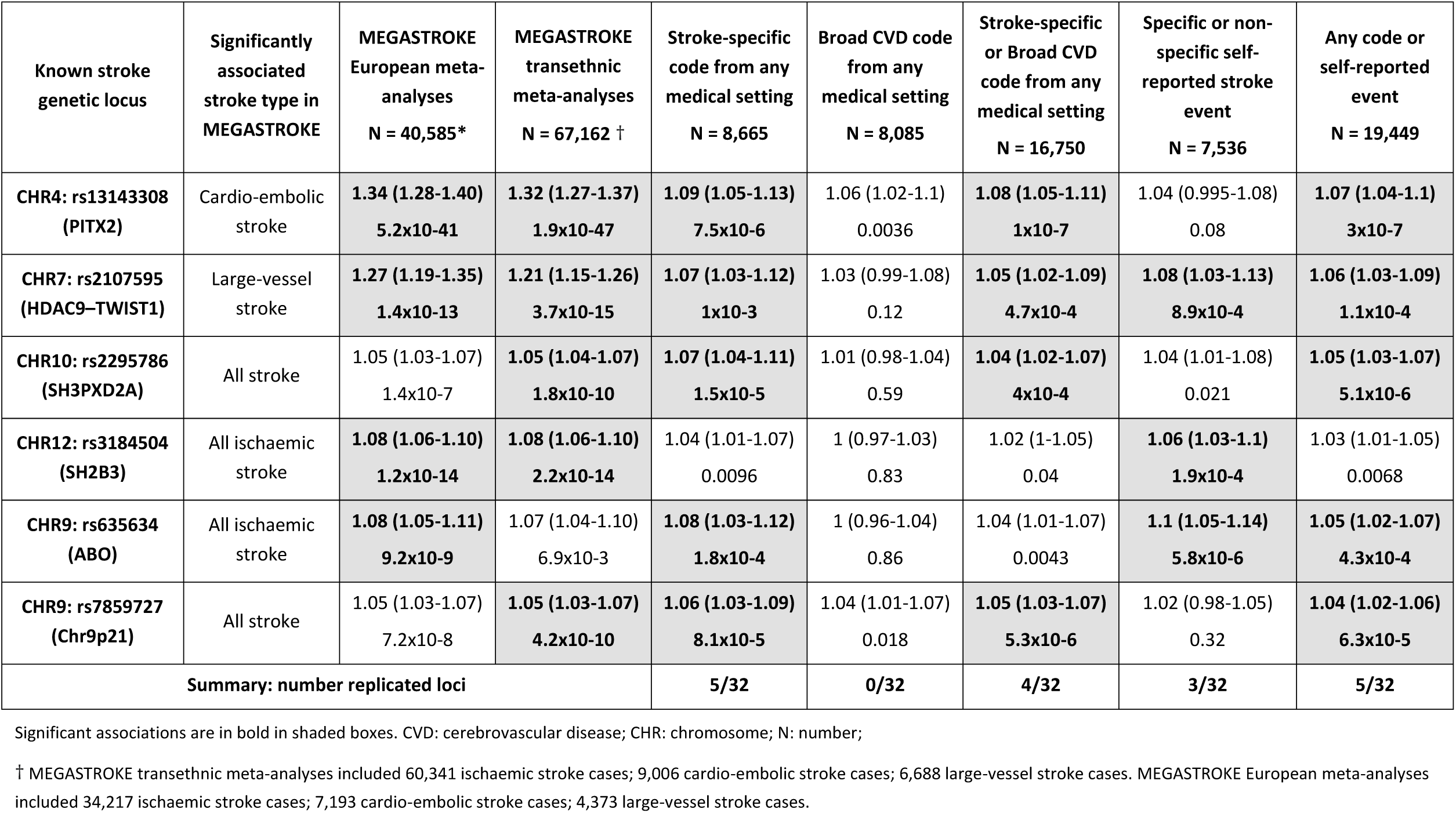
MEGASTROKE stroke-significant loci replicated using UKB stroke definitions.

Power calculations suggested we had ≥80% power to replicate all 32 loci for the any code or self-reported event definition, while having ≥80% power for only 11/32 loci for the definition including a stroke-specific code from any medical setting. (S7 Table).

## Discussion

Our analyses show, that depending on the code source and type used for identifying stroke cases in the UKB, the currently achieved maximum case-sample size can range from ∼4,000 to ∼20,000 – a remarkable 5-fold difference. We go on to demonstrate, that regardless of the code source and type used, the resulting GWAS summary results are significantly genetically correlated with the largest stroke GWAS to date. Finally, when we try to replicate known stroke-significant loci in our data, both stroke-specific codes from any medical setting as well as a broad definition including any code or self-reported event, replicate 5 of the 32 loci. Replication generated broadly similar effect sizes for all but 2 stroke subtype specific loci, which is likely explained by our dataset including a mix of stroke subtypes. Another possible explanation is the “winner’s curse” phenomenon (i.e. the estimated effect of a marker allele from the initial study reporting the marker-allele association is often exaggerated relative to the estimated effect in follow-up studies).

The correlation of all definitions with the MEGASTROKE study results suggests one or more of the following: (i) all definitions retrieve true-positive acute stroke cases, meaning that broad CVD codes include additional true-positive cases not identified by stroke-specific codes; (ii) cases coded with a broad CVD code have not necessarily suffered an acute stroke, but represent a range of phenotypes with a similar genetic architecture to acute stroke (e.g. previous research has shown at least one overlapping locus for carotid artery disease and acute stroke (Malik, 2018)); (iii) the MEGASTROKE study includes some misclassified broad CVD cases as false-positive acute stroke cases. It is most likely that a combination of these factors is contributing to our findings, but we are unable to dissect their separate contributions in the current study.

Previous case-note validation studies suggested that broad CVD codes are better at identifying the broad conditions they signify as opposed to ascertaining acute stroke cases (McCormick, 2015), supporting a role for option two above. An example of this would be a case-note review of patients with a code for an unruptured intracranial aneurysm or carotid artery stenosis confirming that the diagnosis was also most likely an unruptured intracranial aneurysm or carotid artery stenosis, rather than rather than the reviewing clinician deciding it was an acute stroke that had been miscoded as an unruptured intracranial aneurysm or carotid artery stenosis.

Despite the definition using any code or self-reported event increasing the sample size by more than two-fold compared to the definition using only stroke-specific codes from a medical setting, it did not replicate a higher number of known stroke-associated loci. This could suggest that there is still insufficient power to replicate additional loci using any of our definitions despite power calculations suggesting ≥80% power for all loci. Also, associations for nine of the 32 loci in MEGASTROKE were only found for specific stroke subtypes and 11 of the 32 loci were significant in analyses including only ischaemic stroke cases, the proportions of which are unlikely to be identical between the two datasets. For example, the MEGASTROKE study sample included 90% confirmed ischaemic stroke cases. The stroke subtype breakdown among the UKB participants is available only for stroke-specific codes from hospital, death record and self-reported data (UK Biobank datafields ‘42009’, ‘42011’ and ‘42013’) and shows a proportion of confirmed ischaemic stroke cases of 47%, with 10% cases being intracerebral haemorrhage and 11% subarachnoid haemorrhage and the remainder of unspecified stroke subtype. Alternatively, it could also suggest that the additional cases identified by using any code or self-report are not true-positive stroke cases or that some of these known stroke-associated loci are false-positive findings.

Self-reported cases (both stroke-specific and broad CVD) also showed a close genetic correlation with the MEGASTROKE study, supporting the use of self-report as a means of identifying additional stroke cases in the UKB. This was so despite the highly variable results from previous case-note based validation studies of self-report for ascertaining stroke cases. Studying this by case-note validation in UKB itself would be challenging, given the difficulties accessing NHS records which predate recruitment by many years and the fact that participants may have moved between UK regions during their life-course. Other studies have also reported a close genetic correlation between a wide range of self-reported diseases and medical setting diagnoses. Examples include both acute and chronic conditions (e.g. depression, myocardial infarction, rheumatoid arthritis) (Wray, 2018; Howard, 2019; DeBoever, 2020).

For some of our definitions, the rg was >1. The estimated rg is a combination of the true rg and variation. When the true rg is close to the boundary (−1 or 1) and/or variation is large, the estimated rg can go beyond the boundary (Ning, 2020). In rg estimation, some common reasons for generating large variation are: (i) at least one of the h^2^ estimates is very low; (ii) small sample size; (iii) many SNPs in the reference panel are absent in one of the two GWASs; (iv) there is a severe mismatch between the GWAS population and the population for computing reference panel. We can exclude the last two options, and the small sample size is therefore the likely explanation.

In our analyses, the case-control groups were partially overlapping and a specific UKB participant could therefore be a stroke case for one definition and a control for another definition. We used this study design to mimic the ‘real world’ situation, creating binary case-control definitions based on each code list. In theory this could reduce the power of some of the analyses, since it means controls can end up including some true-positive stroke cases. However, in reality it is unlikely to have a significant effect given the overall large number of controls. For example, for analyses using stroke-specific codes from any medical setting, just over 2% of controls have a broad CVD code and/or have self-reported a stroke event.

We used BoltLMM for running the GWAS (Loh, 2015). Our case-fraction ranged from 1% to 5% depending on the case-definition and we limited our analyses to SNPs with a minor allele frequency of at least 1%. Based on simulations done using BoltLMM, the authors of the software suggest that with these case-fraction and minor allele frequency parameters, they did not find a statistically significant inflation of type I error rates (Supplementary table 8 in Loh, 2015).

The strengths of our study are: (i) we included - and have made available to re-use - a clinically informed, comprehensive set of codes across all relevant coding systems; (ii) we compared our results against the largest stroke GWAS to date; (iii) we used multiple methods for comparison accounting for both GWAS significant loci but also SNPs across the whole genome – i.e. correlation and replication; (iv) we have added novel data to what is already known from case-note validation.

Our study also has some limitations: (i) some of our definitions included relatively small case numbers compared to the MEGASTROKE study, reducing our power to replicate known loci; (ii) uneven numbers across definitions not allowing direct comparisons, but rather reflecting the real-world situation; (iii) our definitions included the subarachnoid haemorrhage stroke subtype codes, whereas the MEGASTROKE study did not, resulting in a slightly different mix of stroke cases; (iv) the UKB participants’ demographic characteristics differ from those of the UK general population with evidence of a healthy-volunteer selection bias, which needs to be considered when extrapolating these results to other settings (Fry, 2017).

We have shown that the selection of codes and code sources used to ascertain stroke cases has a major impact on the overall stroke case numbers in the UKB. Given the close genetic correlation between stroke cases identified using broad CVD codes, self-report, and physician-confirmed stroke cases, we suggest that for studies accepting of more crude stroke and cerebrovascular disease outcomes, researchers may wish to include all codes and self-reported events for increased power. Alternatively, this information is also helpful in informing the selection of controls for various studies. Including a large number of broad CVD coded cases among controls might weaken any association seen for certain study designs. However, since we cannot exclude the effects of a shared genetic control of broad CVD phenotypes and acute stroke, this evidence is not sufficient to support using broad CVD codes in studies that need to define acute stroke outcomes very accurately (e.g. clinical trials).

Further research is needed: to better understand the underlying reasons for the close genetic correlation between stroke-specific and broad CVD codes; to dissect the underlying explanation for our results with targeted case-note validation; and to replicate our results in other datasets. In addition, more data is needed on the accuracy of different coding systems for identifying specific pathological stroke subtypes (ischaemic stroke versus intracerebral haemorrhage versus subarachnoid haemorrhage) and aetiological stroke subtypes (e.g., small vessel disease versus large artery disease versus cardioembolic stroke versus other / unknown cause).

## Data Availability

The datasets analyzed for this study is available from the UK Biobank and the MEGASTROKE consortium website upon request (https://www.megastroke.org/).

## Acknowledgements

This work was undertaken under a UKB project 2532 “UK Biobank Stroke Study (UKBiSS): developing an in-depth understanding of the determinants of stroke and its subtypes.” Authors acknowledge Dr Spiros Denaxas, UCL, for his valuable comments on the manuscript. The MEGASTROKE project received funding from sources specified at http://www.megastroke.org/acknowledgments.html and the author list for the MEGASTROKE consortium is added in Appendix 1.

## Author contributions

All authors made substantial contributions to the conception or design of the work; or the acquisition, analysis, or interpretation of data for the work; AND drafting the work or revising it critically for important intellectual content; AND final approval of the version to be published; AND agreement to be accountable for all aspects of the work in ensuring that questions related to the accuracy or integrity of any part of the work are appropriately investigated and resolved.

## Data sharing

The data that supports the findings of this study are available in the supplementary material of this article.

## References

1. Sudlow, C. L. M., Gallacher, J., Allen, N., Beral, V., Burton, B., Danesh, J. et al. (2015). UK Biobank: an open access resource for identifying the causes of a wide range of complex diseases of middle and old age. PLoS Medicine. 31;12(3):e1001779.

2. Lozano, R., Naghavi, M., Foreman, K., Lim, S., Shibuya, K., Aboyans, V. et al. (2010). Global and regional mortality from 235 causes of death for 20 age groups in 1990 and 2010: a systematic analysis for the Global Burden of Disease Study 2010. Lancet. 380(9859):2095–2128.

3. Woodfield, R., Grant, I., UK Biobank Stroke Outcomes group, UK Biobank Follow-Up and Outcomes Working Group, Sudlow, C. L. M. (2015). Accuracy of Electronic Health Record Data for Identifying Stroke Cases in Large-Scale Epidemiological Studies: A Systematic Review from the UK Biobank Stroke Outcomes Group. PLoS One. 10(10):e0140533.

4. Rannikmäe, K., Ngoh, K., Bush, K., Al-Shahi Salman, R., Doubal, F., Flaig, R. et al. (2020). Accuracy of identifying incident stroke cases from linked health care data in UK Biobank. Neurology. 95(6):e697–e707.

5. Woodfield, R., UK Biobank Stroke Outcomes Group, UK Biobank Follow-up and Outcomes Working Group, Sudlow, C. L. M. (2015). Accuracy of Patient Self-Report of Stroke: A Systematic Review from the UK Biobank Stroke Outcomes Group. Plos One. 10(9):e0137538.

6. Liberman, A. L., Rostanski, S. K., Ruff, I. M., Meyer, A. N. D., Maas, M. B., Prabhakaran, S. (2018). Inter-rater Agreement for the Diagnosis of Stroke Versus Stroke Mimic. Neurologist. 23(4):118–121.

7. Manolio, T.A., Goodhand, P., Ginsburg, G. (2020). The International Hundred Thousand Plus Cohort Consortium: integrating large-scale cohorts to address global scientific challenges. Lancet Digital Health. 2(11):e567–e568.

8. Loh, P-R., Tucker, G., Bulik-Sullivan, B. K., Vilhjálmsson, B. J., Finucane, H. K., Salem, R. M. et al. (2015). Efficient Bayesian mixed-model analysis increases association power in large cohorts. Nature Genetics. 47(3):284–290.

9. Lloyd-Jones, L. R., Robinson, M.R., Yang, J., Visscher, P. M. (2018). Transformation of Summary Statistics from Linear Mixed Model Association on All-or-None Traits to Odds Ratio. Genetics. 208(4):1397–1408.

10. Malik, R., Chauhan, G., Traylor, M., Sargurupremraj, M., Okada, Y., Mishra, A. et al. (2018). Multiancestry genome-wide association study of 520,000 subjects identifies 32 loci associated with stroke and stroke subtypes. Nature Genetics. 50(4):524–537.

11. Ning, Z., Pawitan, Y., Shen, X. (2020). High-definition likelihood inference of genetic correlations across human complex traits. Nature Genetics. 52:859–864.

12. Virani, S. S., Alonso, A., Benjamin, E. J., Bittencourt, M. S., Callaway, C. W., Carson, A.P. et al. (2020). Heart Disease and Stroke Statistics - 2020 Update: A Report From the American Heart Association. Circulation. 141(9):e139.

13. McCormick, N., Bhole, V., Lacaille, D., Avina-Zubieta, J. A. (2015). Validity of Diagnostic Codes for Acute Stroke in Administrative Databases: A Systematic Review. PLoS One. 10(8):e0135834.

14. Wray, N. R., Ripke, S., Mattheien, M., Trzaskowski, M., Byrne, E. M., Abdellaoui, A. et al. (2018). Genome-wide association analyses identify 44 risk variants and refine the genetic architecture of major depression. Nature Genetics. 50:668–681.

15. Howard, D. M., Adams, M. J., Clarke, T. K., Hafferty, J. D., Gibson, J., Shirali, M. et al. (2019). Genome-wide meta-analysis of depression identifies 102 independent variants and highlights the importance of the prefrontal brain regions. Nature Neuroscience. 22:343–352.

16. DeBoever, C., Tanigawa, Y., Aguirre, M., McInnes, G., Lavertu, A., Rivas, M. A. (2020). Assessing Digital Phenotyping to Enhance Genetic Studies of Human Diseases. American Journal of Human Genetics. 106(5):611–622.

17. Fry, A., Littlejohns, T. J., Sudlow, C. L. M., Doherty, N., Adamska, L., Sprosen, T. et al. (2017). Comparison of Sociodemographic and Health-Related Characteristics of UK Biobank Participants With Those of the General Population. American Journal of Epidemiology. 186(9):1026–1034.

