## Supplemental Figures and Appendix 1 for "Physician-confirmed and administrative definitions of stroke in UK Biobank reflect the same underlying genetic trait"

**Figure S1. Code sources.**

1a. Code sources for cases with a stroke-specific code

2b. Code sources for cases with a broad cerebrovascular disease code

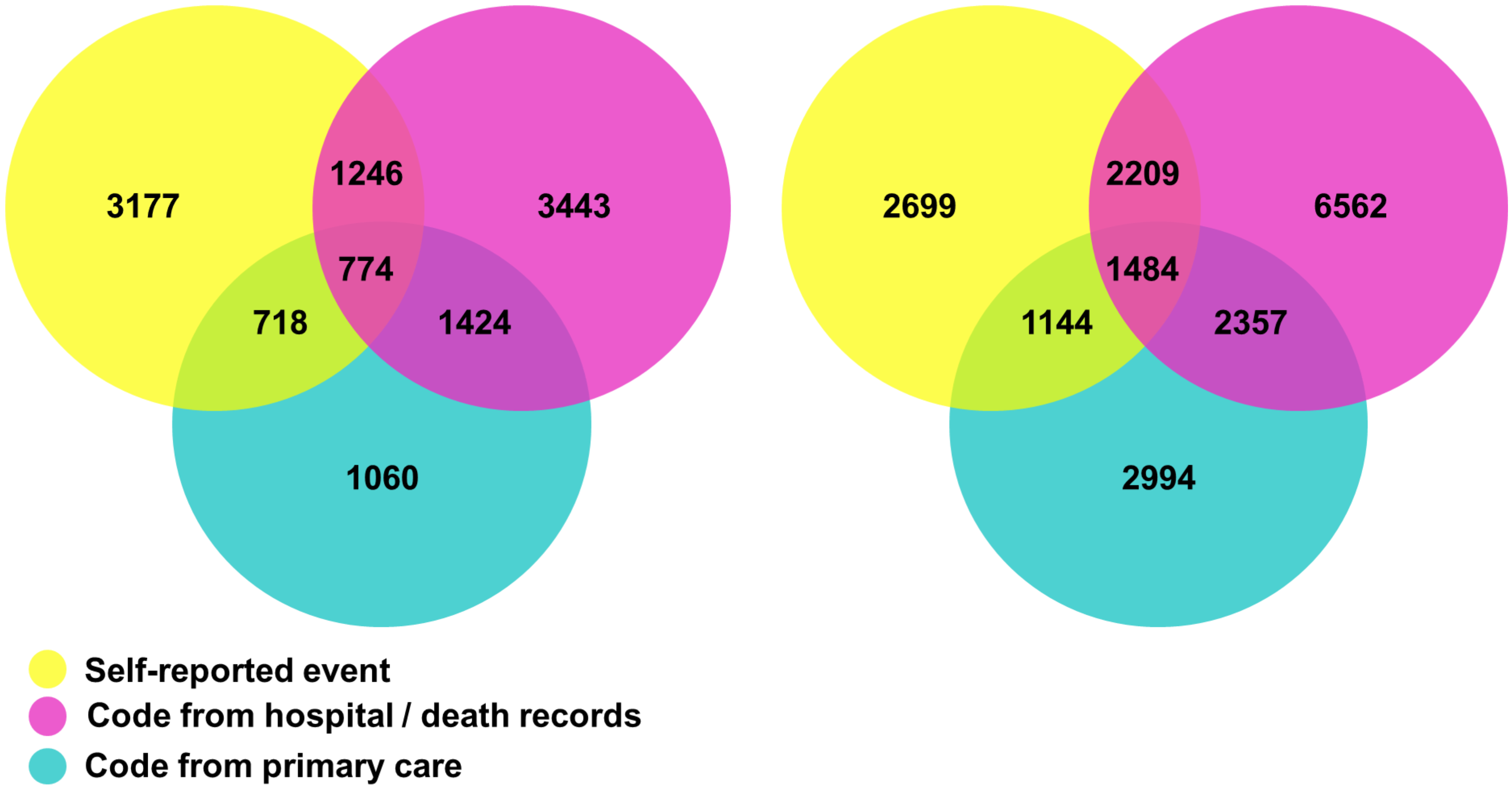

### Figure S2. MEGASTROKE stroke-significant loci replicated using UK Biobank stroke definitions

MEGASTROKE odds ratio and p-value is shown for the analyses (European or trans-ethnic) showing the lowest p-value.

### 2a.

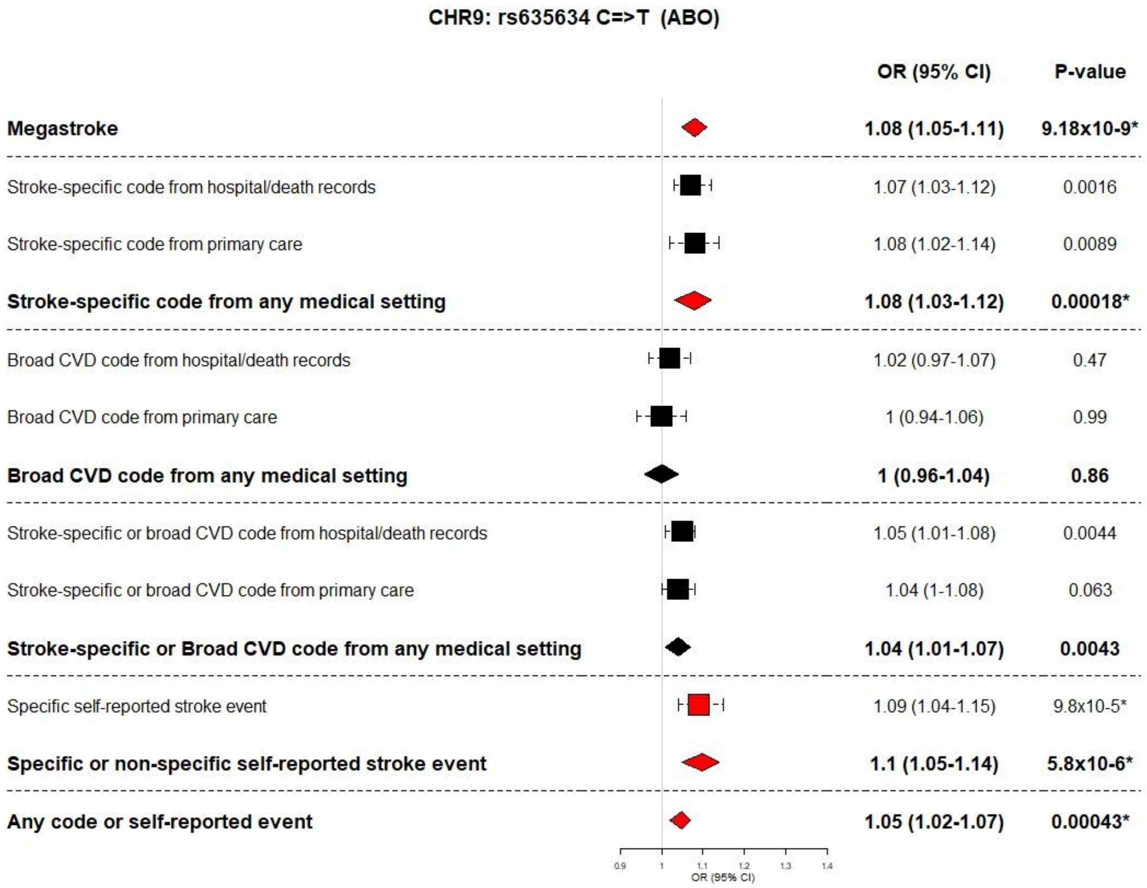

**2b.**

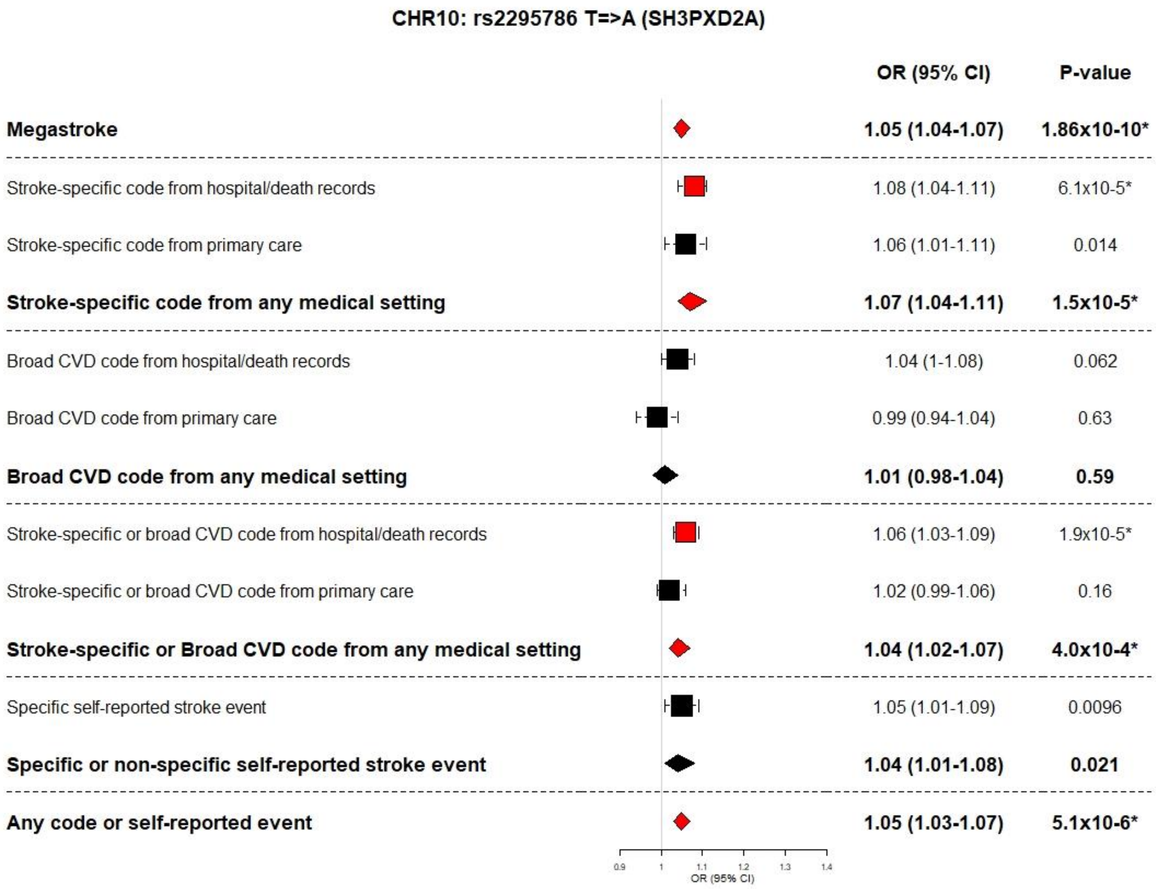

CHR10: rs2295786 T=>A (SH3PXD2A)
OR (95% CI)
P-value
Megastroke
1.05 (1.04-1.07)
1.86x10-10*
Stroke-specific code from hospital/death records
1.08 (1.04-1.11)
6.1x10-5*
Stroke-specific code from primary care
1.06 (1.01-1.11)
0.014
Stroke-specific code from any medical setting
1.07 (1.04-1.11)
1.5x10-5*
Broad CVD code from hospital/death records
1.04 (1-1.08)
0.062
Broad CVD code from primary care
0.99 (0.94-1.04)
0.63
Broad CVD code from any medical setting
1.01 (0.98-1.04)
0.59
Stroke-specific or broad CVD code from hospital/death records
1.06 (1.03-1.09)
1.9x10-5*
Stroke-specific or broad CVD code from primary care
1.02 (0.99-1.06)
0.16
Stroke-specific or Broad CVD code from any medical setting
1.04 (1.02-1.07)
4.0x10-4*
Specific self-reported stroke event
1.05 (1.01-1.09)
0.0096
Specific or non-specific self-reported stroke event
1.04 (1.01-1.08)
0.021
Any code or self-reported event
1.05 (1.03-1.07)
5.1x10-6*
0.9
1
1.1
1.2
1.3
1.4
OR (95% CI)

**2c.**

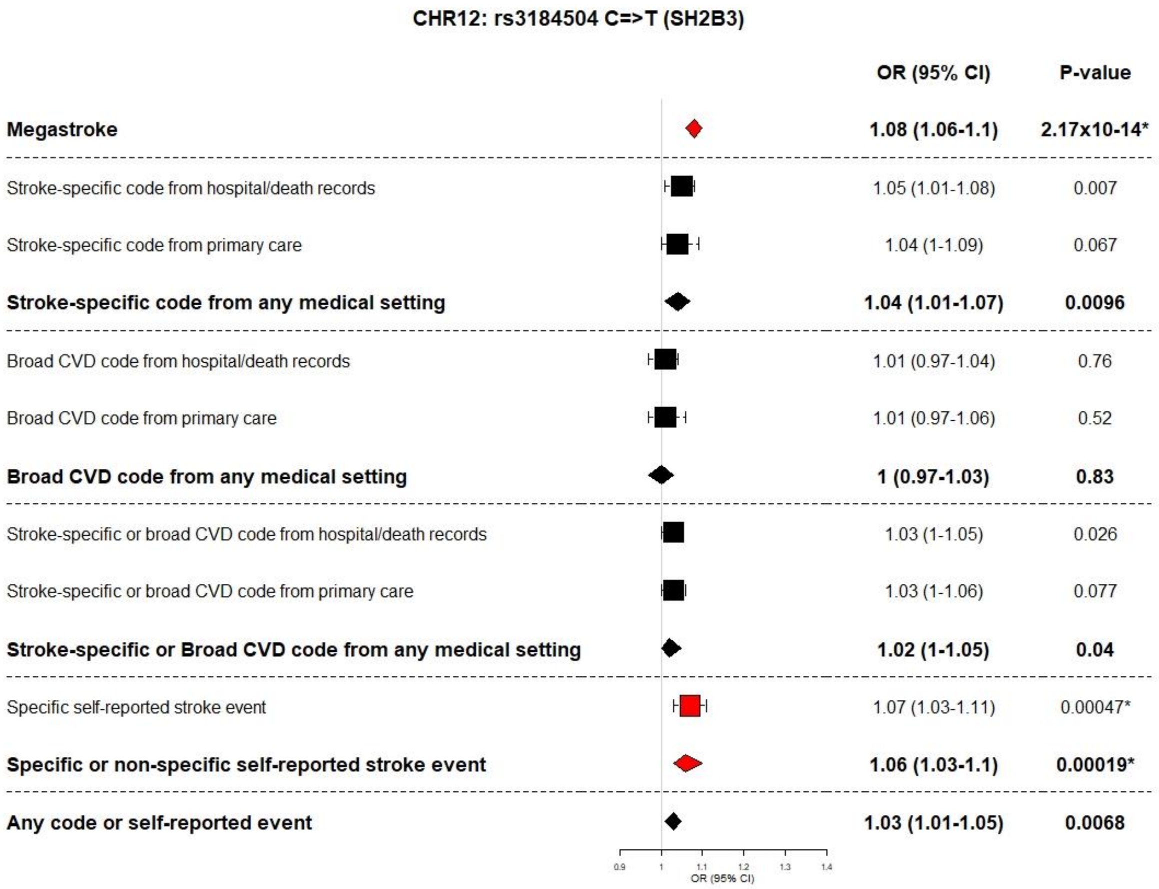
CHR12: rs3184504 C=>T (SH2B3)
OR (95% CI)
P-value
Megastroke
1.08 (1.06-1.1)
2.17x10-14*
Stroke-specific code from hospital/death records
1.05 (1.01-1.08)
0.007
Stroke-specific code from primary care
1.04 (1-1.09)
0.067
Stroke-specific code from any medical setting
1.04 (1.01-1.07)
0.0096
Broad CVD code from hospital/death records
1.01 (0.97-1.04)
0.76
Broad CVD code from primary care
1.01 (0.97-1.06)
0.52
Broad CVD code from any medical setting
1 (0.97-1.03)
0.83
Stroke-specific or broad CVD code from hospital/death records
1.03 (1-1.05)
0.026
Stroke-specific or broad CVD code from primary care
1.03 (1-1.06)
0.077
Stroke-specific or Broad CVD code from any medical setting
1.02 (1-1.05)
0.04
Specific self-reported stroke event
1.07 (1.03-1.11)
0.00047*
Specific or non-specific self-reported stroke event
1.06 (1.03-1.1)
0.00019*
Any code or self-reported event
1.03 (1.01-1.05)
0.0068
0.9
1
1.1
1.2
1.3
1.4
OR (95% CI)

**2d.**

CHR9: rs7859727 C=>T (Chr9p21)

| | OR (95% CI) | P-value |
|---|---|---|
| **Megastroke** | **1.05 (1.03-1.07)** | **4.22x10-10*** |
| Stroke-specific code from hospital/death records | 1.07 (1.04-1.11) | 2.7x10-5* |
| Stroke-specific code from primary care | 1.06 (1.01-1.11) | 0.0091 |
| **Stroke-specific code from any medical setting** | **1.06 (1.03-1.09)** | **8.1x10-5*** |
| Broad CVD code from hospital/death records | 1.04 (1-1.08) | 0.032 |
| Broad CVD code from primary care | 1.04 (0.99-1.08) | 0.11 |
| **Broad CVD code from any medical setting** | **1.04 (1.01-1.07)** | **0.018** |
| Stroke-specific or broad CVD code from hospital/death records | 1.06 (1.03-1.09) | 4.5x10-6* |
| Stroke-specific or broad CVD code from primary care | 1.05 (1.02-1.08) | 0.0027 |
| **Stroke-specific or Broad CVD code from any medical setting** | **1.05 (1.03-1.07)** | **5.3x10-6*** |
| Specific self-reported stroke event | 1.01 (0.97-1.05) | 0.62 |
| **Specific or non-specific self-reported stroke event** | **1.02 (0.98-1.05)** | **0.32** |
| **Any code or self-reported event** | **1.04 (1.02-1.06)** | **6.3x10-5*** |

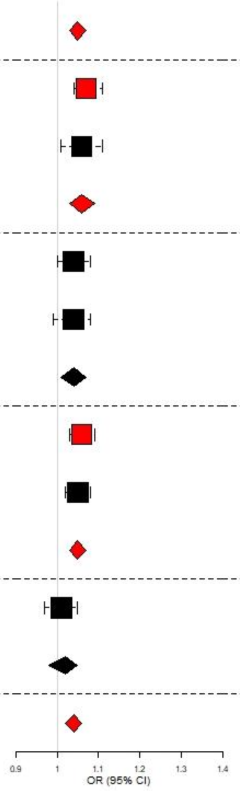

### Appendix 1

**MEGASTROKE CONSORTIUM**

Rainer Malik [1], Ganesh Chauhan [2], Matthew Traylor [3], Muralidharan Sargurupremraj [4,5], Yukinori Okada [6,7,8], Aniket Mishra [4,5], Loes Rutten-Jacobs [3], Anne-Katrin Giese [9], Sander W van der Laan [10], Solveig Gretarsdottir [11], Christopher D Anderson [12,13,14,14], Michael Chong [15], Hieab HH Adams [16,17], Tetsuro Ago [18], Peter Almgren [19], Philippe Amouyel [20,21], Hakan Ay [22,13], Traci M Bartz [23], Oscar R Benavente [24], Steve Bevan [25], Giorgio B Boncoraglio [26], Robert D Brown, Jr. [27], Adam S Butterworth [28,29], Caty Carrera [30,31], Cara L Carty [32,33], Daniel I Chasman [34,35], Wei-Min Chen [36], John W Cole [37], Adolfo Correa [38], Ioana Cotlarciuc [39], Carlos Cruchaga [40,41], John Danesh [28,42,43,44], Paul IW de Bakker [45,46], Anita L DeStefano [47,48], Marcel den Hoed [49], Qing Duan [50], Stefan T Engelter [51,52], Guido J Falcone [53,54], Rebecca F Gottesman [55], Raji P Grewal [56], Vilmundur Gudnason [57,58], Stefan Gustafsson [59], Jeffrey Haessler [60], Tamara B Harris [61], Ahamad Hassan [62], Aki S Havulinna [63,64], Susan R Heckbert [65], Elizabeth G Holliday [66,67], George Howard [68], Fang-Chi Hsu [69], Hyacinth I Hyacinth [70], M Arfan Ikram [16], Erik Ingelsson [71,72], Marguerite R Irvin [73], Xueqiu Jian [74], Jordi Jiménez-Conde [75], Julie A Johnson [76,77], J Wouter Jukema [78], Masahiro Kanai [6,7,79], Keith L Keene [80,81], Brett M Kissela [82], Dawn O Kleindorfer [82], Charles Kooperberg [60], Michiaki Kubo [83], Leslie A Lange [84], Carl D Langefeld [85], Claudia Langenberg [86], Lenore J Launer [87], Jin-Moo Lee [88], Robin Lemmens [89,90], Didier Leys [91], Cathryn M Lewis [92,93], Wei-Yu Lin [28,94], Arne G Lindgren [95,96], Erik Lorentzen [97], Patrik K Magnusson [98], Jane Maguire [99], Ani Manichaikul [36], Patrick F McArdle [100], James F Meschia [101], Braxton D Mitchell [100,102], Thomas H Mosley [103,104], Michael A Nalls [105,106], Toshiharu Ninomiya [107], Martin J O'Donnell [15,108], Bruce M Psaty [109,110,111,112], Sara L Pulit [113,45], Kristiina Rannikmäe [114,115], Alexander P Reiner [65,116], Kathryn M Rexrode [117], Kenneth Rice [118], Stephen S Rich [36], Paul M Ridker [34,35], Natalia S Rost [9,13], Peter M Rothwell [119], Jerome I Rotter [120,121], Tatjana Rundek [122], Ralph L Sacco [122], Saori Sakaue [7,123], Michele M Sale [124], Veikko Salomaa [63], Bishwa R Sapkota [125], Reinhold Schmidt [126], Carsten O Schmidt [127], Ulf Schminke [128], Pankaj Sharma [39], Agnieszka Slowik [129], Cathie LM Sudlow [114,115], Christian Tanislav [130], Turgut Tatlisumak [131,132], Kent D Taylor [120,121], Vincent NS Thijs [133,134], Gudmar Thorleifsson [11], Unnur Thorsteinsdottir [11], Steffen Tiedt [1], Stella Trompet [135], Christophe Tzourio [5,136,137], Cornelia M van Duijn [138,139], Matthew Walters [140], Nicholas J Wareham [86], Sylvia Wassertheil-Smoller [141], James G Wilson [142], Kerri L Wiggins [109], Qiong Yang [47], Salim Yusuf [15], Najaf Amin [16], Hugo S Aparicio [185,48], Donna K Arnett [186], John Attia [187], Alexa S Beiser [47,48], Claudine Berr [188], Julie E Buring [34,35], Mariana Bustamante [189], Valeria Caso [190], Yu-Ching Cheng [191], Seung Hoan Choi [192,48], Ayesha Chowhan [185,48], Natalia Cullell [31], Jean-François Dartigues [193,194], Hossein Delavaran [95,96], Pilar Delgado [195], Marcus Dörr [196,197], Gunnar Engström [19], Ian Ford [198], Wander S Gurpreet [199], Anders Hamsten [200,201], Laura Heitsch [202], Atsushi Hozawa [203], Laura Ibanez [204], Andreea Ilinca [95,96], Martin Ingelsson [205], Motoki Iwasaki [206], Rebecca D Jackson [207], Katarina Jood [208], Pekka Jousilahti [63], Sara Kaffashian [4,5], Lalit Kalra [209], Masahiro Kamouchi [210], Takanari Kitazono [211], Olafur Kjartansson [212], Manja Kloss [213], Peter J Koudstaal [214], Jerzy Krupinski [215], Daniel L Labovitz [216], Cathy C Laurie [118], Christopher R Levi [217], Linxin Li [218], Lars Lind [219], Cecilia M Lindgren [220,221], Vasileios Lioutas [222,48], Yong Mei Liu [223], Oscar L Lopez [224], Hirata Makoto [225], Nicolas Martinez-Majander [172], Koichi Matsuda [225], Naoko Minegishi [203], Joan Montaner [226], Andrew P Morris [227,228], Elena Muiño [31], Martina Müller-Nurasyid [229,230,231], Bo Norrving [95,96], Soichi Ogishima [203], Eugenio A Parati [232], Leema Reddy Peddareddygari [56], Nancy L Pedersen [98,233], Joanna Pera [129], Markus Perola [63,234], Alessandro Pezzini [235], Silvana Pileggi [236], Raquel Rabionet [237], Iolanda Riba-Llena [30], Marta Ribasés [238], Jose R Romero [185,48], Jaume Roquer [239,240], Anthony G Rudd [241,242], Antti-Pekka Sarin [243,244], Ralhan Sarju [199], Chloe Sarnowski [47,48], Makoto Sasaki [245], Claudia L Satizabal [185,48], Mamoru Satoh [245], Naveed Sattar [246], Norie Sawada [206], Gerli Sibolt [172], Ásgeir Sigurdsson [247], Albert Smith [248], Kenji Sobue [245], Carolina Soriano-Tárraga [240], Tara Stanne [249], O Colin Stine [250], David J Stott [251], Konstantin Strauch [229,252], Takako Takai [203], Hideo Tanaka [253,254], Kozo Tanno [245], Alexander Teumer [255], Liisa Tomppo [172], Nuria P Torres-Aguila [31], Emmanuel Touze [256,257], Shoichiro Tsugane [206], Andre G Uitterlinden [258], Einar M Valdimarsson [259], Sven J van der Lee [16], Henry Völzke [255], Kenji Wakai [253], David Weir [260], Stephen R Williams [261], Charles DA Wolfe [241,242], Quenna Wong [118], Huichun Xu [191], Taiki Yamaji [206], Dharambir K Sanghera [125,169,170], Olle

Melander [19], Christina Jern [171], Daniel Strbian [172,173], Israel Fernandez-Cadenas [31,30], W T Longstreth, Jr [174,65], Arndt Rolfs [175], Jun Hata [107], Daniel Woo [82], Jonathan Rosand [12,13,14], Guillaume Pare [15], Jemma C Hopewell [176], Danish Saleheen [177], Kari Stefansson [11,178], Bradford B Worrall [179], Steven J Kittner [37], Sudha Seshadri [180,48], Myriam Fornage [74,181], Hugh S Markus [3], Joanna MM Howson [28], Yoichiro Kamatani [6,182], Stephanie Debette [4,5], Martin Dichgans [1,183,184]

1 Institute for Stroke and Dementia Research (ISD), University Hospital, LMU Munich, Munich, Germany
2 Centre for Brain Research, Indian Institute of Science, Bangalore, India
3 Stroke Research Group, Division of Clinical Neurosciences, University of Cambridge, UK
4 INSERM U1219 Bordeaux Population Health Research Center, Bordeaux, France
5 University of Bordeaux, Bordeaux, France
6 Laboratory for Statistical Analysis, RIKEN Center for Integrative Medical Sciences, Yokohama, Japan
7 Department of Statistical Genetics, Osaka University Graduate School of Medicine, Osaka, Japan
8 Laboratory of Statistical Immunology, Immunology Frontier Research Center (WPI-IFReC), Osaka University, Suita, Japan.
9 Department of Neurology, Massachusetts General Hospital, Harvard Medical School, Boston, MA, USA
10 Laboratory of Experimental Cardiology, Division of Heart and Lungs, University Medical Center Utrecht, University of Utrecht, Utrecht,Netherlands
11 deCODE genetics/AMGEN inc, Reykjavik, Iceland
12 Center for Genomic Medicine, Massachusetts General Hospital (MGH), Boston, MA, USA
13 J. Philip Kistler Stroke Research Center, Department of Neurology, MGH, Boston, MA, USA
14 Program in Medical and Population Genetics, Broad Institute, Cambridge, MA, USA
15 Population Health Research Institute, McMaster University, Hamilton, Canada
16 Department of Epidemiology, Erasmus University Medical Center, Rotterdam, Netherlands
17 Department of Radiology and Nuclear Medicine, Erasmus University Medical Center, Rotterdam, Netherlands
18 Department of Medicine and Clinical Science, Graduate School of Medical Sciences, Kyushu University, Fukuoka, Japan
19 Department of Clinical Sciences, Lund University, Malmö, Sweden
20 Univ. Lille, Inserm, Institut Pasteur de Lille, LabEx DISTALZ-UMR1167, Risk factors and molecular determinants of aging-related diseases, F-59000 Lille, France
21 Centre Hosp. Univ Lille, Epidemiology and Public Health Department, F-59000 Lille, France
22 AA Martinos Center for Biomedical Imaging, Department of Radiology, Massachusetts General Hospital, Harvard Medical School, Boston, MA, USA
23 Cardiovascular Health Research Unit, Departments of Biostatistics and Medicine, University of Washington, Seattle, WA, USA
24 Division of Neurology, Faculty of Medicine, Brain Research Center, University of British Columbia, Vancouver, Canada
25 School of Life Science, University of Lincoln, Lincoln, UK
26 Department of Cerebrovascular Diseases, Fondazione IRCCS Istituto Neurologico "Carlo Besta", Milano, Italy
27 Department of Neurology, Mayo Clinic Rochester, Rochester, MN, USA
28 MRC/BHF Cardiovascular Epidemiology Unit, Department of Public Health and Primary Care, University of Cambridge, Cambridge, UK
29 The National Institute for Health Research Blood and Transplant Research Unit in Donor Health and Genomics, University of Cambridge, UK
30 Neurovascular Research Laboratory, Vall d'Hebron Institut of Research, Neurology and Medicine Departments-Universitat Autònoma de Barcelona, Vall d'Hebrón Hospital, Barcelona, Spain
31 Stroke Pharmacogenomics and Genetics, Fundacio Docència i Recerca MutuaTerrassa, Terrassa, Spain
32 Children's Research Institute, Children's National Medical Center, Washington, DC, USA
33 Center for Translational Science, George Washington University, Washington, DC, USA
34 Division of Preventive Medicine, Brigham and Women's Hospital, Boston, MA, USA

35 Harvard Medical School, Boston, MA, USA
36 Center for Public Health Genomics, Department of Public Health Sciences, University of Virginia, Charlottesville, VA, USA
37 Department of Neurology, University of Maryland School of Medicine and Baltimore VAMC, Baltimore, MD, USA
38 Departments of Medicine, Pediatrics and Population Health Science, University of Mississippi Medical Center, Jackson, MS, USA
39 Institute of Cardiovascular Research, Royal Holloway University of London, UK & Ashford and St Peters Hospital, Surrey UK
40 Department of Psychiatry,The Hope Center Program on Protein Aggregation and Neurodegeneration (HPAN),Washington University, School of Medicine, St. Louis, MO, USA
41 Department of Developmental Biology, Washington University School of Medicine, St. Louis, MO, USA
42 NIHR Blood and Transplant Research Unit in Donor Health and Genomics, Department of Public Health and Primary Care, University of Cambridge, Cambridge, UK
43 Wellcome Trust Sanger Institute, Wellcome Trust Genome Campus, Hinxton, Cambridge, UK
44 British Heart Foundation, Cambridge Centre of Excellence, Department of Medicine, University of Cambridge, Cambridge, UK
45 Department of Medical Genetics, University Medical Center Utrecht, Utrecht, Netherlands
46 Department of Epidemiology, Julius Center for Health Sciences and Primary Care, University Medical Center Utrecht, Utrecht, Netherlands
47 Boston University School of Public Health, Boston, MA, USA
48 Framingham Heart Study, Framingham, MA, USA
49 Department of Immunology, Genetics and Pathology and Science for Life Laboratory, Uppsala University, Uppsala, Sweden
50 Department of Genetics, University of North Carolina, Chapel Hill, NC, USA
51 Department of Neurology and Stroke Center, Basel University Hospital, Switzerland
52 Neurorehabilitation Unit, University and University Center for Medicine of Aging and Rehabilitation Basel, Felix Platter Hospital, Basel, Switzerland
53 Department of Neurology, Yale University School of Medicine, New Haven, CT, USA
54 Program in Medical and Population Genetics, The Broad Institute of Harvard and MIT, Cambridge, MA, USA
55 Department of Neurology, Johns Hopkins University School of Medicine, Baltimore, MD, USA
56 Neuroscience Institute, SF Medical Center, Trenton, NJ, USA
57 Icelandic Heart Association Research Institute, Kopavogur, Iceland
58 University of Iceland, Faculty of Medicine, Reykjavik, Iceland
59 Department of Medical Sciences, Molecular Epidemiology and Science for Life Laboratory, Uppsala University, Uppsala, Sweden
60 Division of Public Health Sciences, Fred Hutchinson Cancer Research Center, Seattle, WA, USA
61 Laboratory of Epidemiology and Population Science, National Institute on Aging, National Institutes of Health, Bethesda, MD, USA
62 Department of Neurology, Leeds General Infirmary, Leeds Teaching Hospitals NHS Trust, Leeds, UK
63 National Institute for Health and Welfare, Helsinki, Finland
64 FIMM - Institute for Molecular Medicine Finland, Helsinki, Finland
65 Department of Epidemiology, University of Washington, Seattle, WA, USA
66 Public Health Stream, Hunter Medical Research Institute, New Lambton, Australia
67 Faculty of Health and Medicine, University of Newcastle, Newcastle, Australia
68 School of Public Health, University of Alabama at Birmingham, Birmingham, AL, USA
69 Department of Biostatistical Sciences, Wake Forest School of Medicine, Winston-Salem, NC, USA
70 Aflac Cancer and Blood Disorder Center, Department of Pediatrics, Emory University School of Medicine, Atlanta, GA, USA
71 Department of Medicine, Division of Cardiovascular Medicine, Stanford University School of Medicine, CA, USA

72 Department of Medical Sciences, Molecular Epidemiology and Science for Life Laboratory, Uppsala University, Uppsala, Sweden
73 Epidemiology, School of Public Health, University of Alabama at Birmingham, USA
74 Brown Foundation Institute of Molecular Medicine, University of Texas Health Science Center at Houston, Houston, TX, USA
75 Neurovascular Research Group (NEUVAS), Neurology Department, Institut Hospital del Mar d'Investigació Mèdica, Universitat Autònoma de Barcelona, Barcelona, Spain
76 Department of Pharmacotherapy and Translational Research and Center for Pharmacogenomics, University of Florida, College of Pharmacy, Gainesville, FL, USA
77 Division of Cardiovascular Medicine, College of Medicine, University of Florida, Gainesville, FL, USA
78 Department of Cardiology, Leiden University Medical Center, Leiden, the Netherlands
79 Program in Bioinformatics and Integrative Genomics, Harvard Medical School, Boston, MA, USA
80 Department of Biology, East Carolina University, Greenville, NC, USA
81 Center for Health Disparities, East Carolina University, Greenville, NC, USA
82 University of Cincinnati College of Medicine, Cincinnati, OH, USA
83 RIKEN Center for Integrative Medical Sciences, Yokohama, Japan
84 Department of Medicine, University of Colorado Denver, Anschutz Medical Campus, Aurora, CO, USA
85 Center for Public Health Genomics and Department of Biostatistical Sciences, Wake Forest School of Medicine, Winston-Salem, NC, USA
86 MRC Epidemiology Unit, University of Cambridge School of Clinical Medicine, Institute of Metabolic Science, Cambridge Biomedical Campus, Cambridge, UK
87 Intramural Research Program, National Institute on Aging, National Institutes of Health, Bethesda, MD, USA
88 Department of Neurology, Radiology, and Biomedical Engineering, Washington University School of Medicine, St. Louis, MO, USA
89 KU Leuven – University of Leuven, Department of Neurosciences, Experimental Neurology, Leuven, Belgium
90 VIB Center for Brain & Disease Research, University Hospitals Leuven, Department of Neurology, Leuven, Belgium
91 Univ.-Lille, INSERM U 1171. CHU Lille. Lille, France
92 Department of Medical and Molecular Genetics, King's College London, London, UK
93 SGDP Centre, Institute of Psychiatry, Psychology & Neuroscience, King's College London, London, UK
94 Northern Institute for Cancer Research, Paul O'Gorman Building, Newcastle University, Newcastle, UK
95 Department of Clinical Sciences Lund, Neurology, Lund University, Lund, Sweden
96 Department of Neurology and Rehabilitation Medicine, Skåne University Hospital, Lund, Sweden
97 Bioinformatics Core Facility, University of Gothenburg, Gothenburg, Sweden
98 Department of Medical Epidemiology and Biostatistics, Karolinska Institutet, Stockholm, Sweden
99 University of Technology Sydney, Faculty of Health, Ultimo, Australia
100 Department of Medicine, University of Maryland School of Medicine, MD, USA
101 Department of Neurology, Mayo Clinic, Jacksonville, FL, USA
102 Geriatrics Research and Education Clinical Center, Baltimore Veterans Administration Medical Center, Baltimore, MD, USA
103 Division of Geriatrics, School of Medicine, University of Mississippi Medical Center, Jackson, MS, USA
104 Memory Impairment and Neurodegenerative Dementia Center, University of Mississippi Medical Center, Jackson, MS, USA
105 Laboratory of Neurogenetics, National Institute on Aging, National institutes of Health, Bethesda, MD, USA
106 Data Tecnica International, Glen Echo MD, USA
107 Department of Epidemiology and Public Health, Graduate School of Medical Sciences, Kyushu University, Fukuoka, Japan

108 Clinical Research Facility, Department of Medicine, NUI Galway, Galway, Ireland
109 Cardiovascular Health Research Unit, Department of Medicine, University of Washington, Seattle, WA, USA
110 Department of Epidemiology, University of Washington, Seattle, WA
111 Department of Health Services, University of Washington, Seattle, WA, USA
112 Kaiser Permanente Washington Health Research Institute, Seattle, WA, USA
113 Brain Center Rudolf Magnus, Department of Neurology, University Medical Center Utrecht, Utrecht, The Netherlands
114 Usher Institute of Population Health Sciences and Informatics, University of Edinburgh, Edinburgh, UK
115 Centre for Clinical Brain Sciences, University of Edinburgh, Edinburgh, UK
116 Fred Hutchinson Cancer Research Center, University of Washington, Seattle, WA, USA
117 Department of Medicine, Brigham and Women's Hospital, Boston, MA, USA
118 Department of Biostatistics, University of Washington, Seattle, WA, USA
119 Nuffield Department of Clinical Neurosciences, University of Oxford, UK
120 Institute for Translational Genomics and Population Sciences, Los Angeles Biomedical Research Institute at Harbor-UCLA Medical Center, Torrance, CA, USA
121 Division of Genomic Outcomes, Department of Pediatrics, Harbor-UCLA Medical Center, Torrance, CA, USA
122 Department of Neurology, Miller School of Medicine, University of Miami, Miami, FL, USA
123 Department of Allergy and Rheumatology, Graduate School of Medicine, the University of Tokyo, Tokyo, Japan
124 Center for Public Health Genomics, University of Virginia, Charlottesville, VA, USA
125 Department of Pediatrics, College of Medicine, University of Oklahoma Health Sciences Center, Oklahoma City, OK, USA
126 Department of Neurology, Medical University of Graz, Graz, Austria
127 University Medicine Greifswald, Institute for Community Medicine, SHIP-KEF, Greifswald, Germany
128 University Medicine Greifswald, Department of Neurology, Greifswald, Germany
129 Department of Neurology, Jagiellonian University, Krakow, Poland
130 Department of Neurology, Justus Liebig University, Giessen, Germany
131 Department of Clinical Neurosciences/Neurology, Institute of Neuroscience and Physiology, Sahlgrenska Academy at University of Gothenburg, Gothenburg, Sweden
132 Sahlgrenska University Hospital, Gothenburg, Sweden
133 Stroke Division, Florey Institute of Neuroscience and Mental Health, University of Melbourne, Heidelberg, Australia
134 Austin Health, Department of Neurology, Heidelberg, Australia
135 Department of Internal Medicine, Section Gerontology and Geriatrics, Leiden University Medical Center, Leiden, the Netherlands
136 INSERM U1219, Bordeaux, France
137 Department of Public Health, Bordeaux University Hospital, Bordeaux, France
138 Genetic Epidemiology Unit, Department of Epidemiology, Erasmus University Medical Center Rotterdam, Netherlands
139 Center for Medical Systems Biology, Leiden, Netherlands
140 School of Medicine, Dentistry and Nursing at the University of Glasgow, Glasgow, UK
141 Department of Epidemiology and Population Health, Albert Einstein College of Medicine, NY, USA
142 Department of Physiology and Biophysics, University of Mississippi Medical Center, Jackson, MS, USA
143 A full list of members and affiliations appears in the Supplementary Note
144 Department of Human Genetics, McGill University, Montreal, Canada
145 Department of Pathophysiology, Institute of Biomedicine and Translation Medicine, University of Tartu, Tartu, Estonia
146 Department of Cardiac Surgery, Tartu University Hospital, Tartu, Estonia
147 Clinical Gene Networks AB,Stockholm, Sweden
148 Department of Genetics and Genomic Sciences, The Icahn Institute for Genomics and Multiscale Biology Icahn School of Medicine at Mount Sinai, New York, NY , USA

149 Department of Pathophysiology, Institute of Biomedicine and Translation Medicine, University of Tartu, Biomeedikum, Tartu, Estonia
150 Integrated Cardio Metabolic Centre, Department of Medicine, Karolinska Institutet, Karolinska Universitetssjukhuset, Huddinge, Sweden.
151 Clinical Gene Networks AB, Stockholm, Sweden
152 Sorbonne Universités, UPMC Univ. Paris 06, INSERM, UMR_S 1166, Team Genomics & Pathophysiology of Cardiovascular Diseases, Paris, France
153 ICAN Institute for Cardiometabolism and Nutrition, Paris, France
154 Department of Biomedical Engineering, University of Virginia, Charlottesville, VA, USA
155 Group Health Research Institute, Group Health Cooperative, Seattle, WA, USA
156 Seattle Epidemiologic Research and Information Center, VA Office of Research and Development, Seattle, WA, USA
157 Cardiovascular Research Center, Massachusetts General Hospital, Boston, MA, USA
158 Department of Medical Research, Bærum Hospital, Vestre Viken Hospital Trust, Gjettum, Norway
159 Saw Swee Hock School of Public Health, National University of Singapore and National University Health System, Singapore
160 National Heart and Lung Institute, Imperial College London, London, UK
161 Department of Gene Diagnostics and Therapeutics, Research Institute, National Center for Global Health and Medicine, Tokyo, Japan
162 Department of Epidemiology, Tulane University School of Public Health and Tropical Medicine, New Orleans, LA, USA
163 Department of Cardiology,University Medical Center Groningen, University of Groningen, Netherlands
164 MRC-PHE Centre for Environment and Health, School of Public Health, Department of Epidemiology and Biostatistics, Imperial College London, London, UK
165 Department of Epidemiology and Biostatistics, Imperial College London, London, UK
166 Department of Cardiology, Ealing Hospital NHS Trust, Southall, UK
167 National Heart, Lung and Blood Research Institute, Division of Intramural Research, Population Sciences Branch, Framingham, MA, USA
168 A full list of members and affiliations appears at the end of the manuscript
169 Department of Phamaceutical Sciences, Collge of Pharmacy, University of Oklahoma Health Sciences Center, Oklahoma City, OK, USA
170 Oklahoma Center for Neuroscience, Oklahoma City, OK, USA
171 Department of Pathology and Genetics, Institute of Biomedicine, The Sahlgrenska Academy at University of Gothenburg, Gothenburg, Sweden
172 Department of Neurology, Helsinki University Hospital, Helsinki, Finland
173 Clinical Neurosciences, Neurology, University of Helsinki, Helsinki, Finland
174 Department of Neurology, University of Washington, Seattle, WA, USA
175 Albrecht Kossel Institute, University Clinic of Rostock, Rostock, Germany
176 Clinical Trial Service Unit and Epidemiological Studies Unit, Nuffield Department of Population Health, University of Oxford, Oxford, UK
177 Department of Genetics, Perelman School of Medicine, University of Pennsylvania, PA, USA
178 Faculty of Medicine, University of Iceland, Reykjavik, Iceland
179 Departments of Neurology and Public Health Sciences, University of Virginia School of Medicine, Charlottesville, VA, USA
180 Department of Neurology, Boston University School of Medicine, Boston, MA, USA
181 Human Genetics Center, University of Texas Health Science Center at Houston, Houston, TX, USA
182 Center for Genomic Medicine, Kyoto University Graduate School of Medicine, Kyoto, Japan
183 Munich Cluster for Systems Neurology (SyNergy), Munich, Germany
184 German Center for Neurodegenerative Diseases (DZNE), Munich, Germany
185 Boston University School of Medicine, Boston, MA, USA
186 University of Kentucky College of Public Health, Lexington, KY, USA
187 University of Newcastle and Hunter Medical Research Institute, New Lambton, Australia
188 Univ. Montpellier, Inserm, U1061, Montpellier, France
189 Centre for Research in Environmental Epidemiology, Barcelona, Spain
190 Department of Neurology, Università degli Studi di Perugia, Umbria, Italy

191 Department of Medicine, University of Maryland School of Medicine, Baltimore, MD, USA
192 Broad Institute, Cambridge, MA, USA
193 Univ. Bordeaux, Inserm, Bordeaux Population Health Research Center, UMR 1219, Bordeaux, France
194 Bordeaux University Hospital, Department of Neurology, Memory Clinic, Bordeaux, France
195 Neurovascular Research Laboratory. Vall d'Hebron Institut of Research, Neurology and Medicine Departments-Universitat Autònoma de Barcelona. Vall d'Hebrón Hospital, Barcelona, Spain
196 University Medicine Greifswald, Department of Internal Medicine B, Greifswald, Germany
197 DZHK, Greifswald, Germany
198 Robertson Center for Biostatistics, University of Glasgow, Glasgow, UK
199 Hero DMC Heart Institute, Dayanand Medical College & Hospital, Ludhiana, India
200 Atherosclerosis Research Unit, Department of Medicine Solna, Karolinska Institutet, Stockholm, Sweden
201 Karolinska Institutet, Stockholm, Sweden
202 Division of Emergency Medicine, and Department of Neurology, Washington University School of Medicine, St. Louis, MO, USA
203 Tohoku Medical Megabank Organization, Sendai, Japan
204 Department of Psychiatry, Washington University School of Medicine, St. Louis, MO, USA
205 Department of Public Health and Caring Sciences / Geriatrics, Uppsala University, Uppsala, Sweden
206 Epidemiology and Prevention Group, Center for Public Health Sciences, National Cancer Center, Tokyo, Japan
207 Department of Internal Medicine and the Center for Clinical and Translational Science, The Ohio State University, Columbus, OH, USA
208 Institute of Neuroscience and Physiology, the Sahlgrenska Academy at University of Gothenburg, Goteborg, Sweden
209 Department of Basic and Clinical Neurosciences, King's College London, London, UK
210 Department of Health Care Administration and Management, Graduate School of Medical Sciences, Kyushu University, Japan
211 Department of Medicine and Clinical Science, Graduate School of Medical Sciences, Kyushu University, Japan
212 Landspitali National University Hospital, Departments of Neurology & Radiology, Reykjavik, Iceland
213 Department of Neurology, Heidelberg University Hospital, Germany
214 Department of Neurology, Erasmus University Medical Center
215 Hospital Universitari Mutua Terrassa, Terrassa (Barcelona), Spain
216 Albert Einstein College of Medicine, Montefiore Medical Center, New York, NY, USA
217 John Hunter Hospital, Hunter Medical Research Institute and University of Newcastle, Newcastle, NSW, Australia
218 Centre for Prevention of Stroke and Dementia, Nuffield Department of Clinical Neurosciences, University of Oxford, UK
219 Department of Medical Sciences, Uppsala University, Uppsala, Sweden
220 Genetic and Genomic Epidemiology Unit, Wellcome Trust Centre for Human Genetics, University of Oxford, Oxford, UK
221 The Wellcome Trust Centre for Human Genetics, Oxford, UK
222 Beth Israel Deaconess Medical Center, Boston, MA, USA
223 Wake Forest School of Medicine, Wake Forest, NC, USA
224 Department of Neurology, University of Pittsburgh, Pittsburgh, PA, USA
225 BioBank Japan, Laboratory of Clinical Sequencing, Department of Computational biology and medical Sciences, Graduate school of Frontier Sciences, The University of Tokyo, Tokyo, Japan
226 Neurovascular Research Laboratory, Vall d'Hebron Institut of Research, Neurology and Medicine Departments-Universitat Autònoma de Barcelona. Vall d'Hebrón Hospital, Barcelona, Spain
227 Department of Biostatistics, University of Liverpool, Liverpool, UK
228 Wellcome Trust Centre for Human Genetics, University of Oxford, Oxford, UK

229 Institute of Genetic Epidemiology, Helmholtz Zentrum München - German Research Center for Environmental Health, Neuherberg, Germany
230 Department of Medicine I, Ludwig-Maximilians-Universität, Munich, Germany
231 DZHK (German Centre for Cardiovascular Research), partner site Munich Heart Alliance, Munich, Germany
232 Department of Cerebrovascular Diseases, Fondazione IRCCS Istituto Neurologico "Carlo Besta", Milano, Italy
233 Karolinska Institutet, MEB, Stockholm, Sweden
234 University of Tartu, Estonian Genome Center, Tartu, Estonia, Tartu, Estonia
235 Department of Clinical and Experimental Sciences, Neurology Clinic, University of Brescia, Italy
236 Translational Genomics Unit, Department of Oncology, IRCCS Istituto di Ricerche Farmacologiche Mario Negri, Milano, Italy
237 Department of Genetics, Microbiology and Statistics, University of Barcelona, Barcelona, Spain
238 Psychiatric Genetics Unit, Group of Psychiatry, Mental Health and Addictions, Vall d'Hebron Research Institute (VHIR), Universitat Autònoma de Barcelona, Biomedical Network Research Centre on Mental Health (CIBERSAM), Barcelona, Spain
239 Department of Neurology, IMIM-Hospital del Mar, and Universitat Autònoma de Barcelona, Spain
240 IMIM (Hospital del Mar Medical Research Institute), Barcelona, Spain
241 National Institute for Health Research Comprehensive Biomedical Research Centre, Guy's & St. Thomas' NHS Foundation Trust and King's College London, London, UK
242 Division of Health and Social Care Research, King's College London, London, UK
243 FIMM-Institute for Molecular Medicine Finland, Helsinki, Finland
244 THL-National Institute for Health and Welfare, Helsinki, Finland
245 Iwate Tohoku Medical Megabank Organization, Iwate Medical University, Iwate, Japan
246 BHF Glasgow Cardiovascular Research Centre, Faculty of Medicine, Glasgow, UK
247 deCODE Genetics/Amgen, Inc., Reykjavik, Iceland
248 Icelandic Heart Association, Reykjavik, Iceland
249 Institute of Biomedicine, the Sahlgrenska Academy at University of Gothenburg, Goteborg, Sweden
250 Department of Epidemiology, University of Maryland School of Medicine, Baltimore, MD, USA
251 Institute of Cardiovascular and Medical Sciences, Faculty of Medicine, University of Glasgow, Glasgow, UK
252 Chair of Genetic Epidemiology, IBE, Faculty of Medicine, LMU Munich, Germany
253 Division of Epidemiology and Prevention, Aichi Cancer Center Research Institute, Nagoya, Japan
254 Department of Epidemiology, Nagoya University Graduate School of Medicine, Nagoya, Japan
255 University Medicine Greifswald, Institute for Community Medicine, SHIP-KEF, Greifswald, Germany
256 Department of Neurology, Caen University Hospital, Caen, France
257 University of Caen Normandy, Caen, France
258 Department of Internal Medicine, Erasmus University Medical Center, Rotterdam, Netherlands
259 Landspitali University Hospital, Reykjavik, Iceland
260 Survey Research Center, University of Michigan, Ann Arbor, MI, USA
261 University of Virginia Department of Neurology, Charlottesville, VA, USA
